# Analysis of genetic risk factors for Leber hereditary optic neuropathy in the Polish population

**DOI:** 10.64898/2026.01.14.26343833

**Authors:** Julia Sikorska, Maciej R. Krawczyński, Magdalena Korwin, Monika Ołdak, Ewa Bartnik, Katarzyna Tońska, Agnieszka Piotrowska-Nowak

## Abstract

Leber hereditary optic neuropathy (LHON) is primarily caused by pathogenic mitochondrial DNA (mtDNA) variants, most commonly the m.11778G>A variant in the *MT-ND4* gene. The presence of this variant alone is insufficient to trigger disease symptoms, of which vision loss is the hallmark. Given the incomplete penetrance and inter-population variability in modifying factors, this study aimed to investigate two previously proposed genetic risk factors for LHON in the Polish population. Using quantitative PCR, we measured the mtDNA copy number in peripheral blood of affected and unaffected carriers of the m.11778G>A variant. In addition, we assessed the frequency of the *PRICKLE3* c.157C>T variant in symptomatic, asymptomatic and control individuals using PCR-RFLP. Our results indicate that neither mtDNA copy number nor the presence of the *PRICKLE3* variant is associated with LHON symptom manifestation in the Polish cohort under conditions tested, in contrast to previously reported associations in other populations. These findings suggest that the incomplete penetrance of LHON in the Polish population may involve other modifying factors, such as yet unidentified nuclear DNA variants.

**Research highlights:**

- Mitochondrial DNA (mtDNA) copy number and the presence of the c.157C>T variant in the *PRICKLE3* gene do not influence the manifestation of Leber hereditary optic neuropathy (LHON) symptoms in the Polish population.
- The results support a geographic dependence of genetic risk factors affecting the penetrance of LHON-associated mtDNA variants.

## 1 Introduction

Dysfunction of the oxidative phosphorylation system (OXPHOS) can lead to various mitochondrial diseases, which may occur at any age. These disorders can follow autosomal, X-linked, or maternal inheritance patterns (Leonard and Schapira 2000). Post-mitotic tissues with high-energy demands, such as muscles and nerves, are typically the most affected (McFarland et al. 2010; Chinnery and Hudson 2013). Among mitochondrial diseases caused by primary pathogenic variants in mitochondrial DNA (mtDNA), Leber hereditary optic neuropathy (LHON) is the most common, affecting approximately 1 in 30,000 to 50,000 Europeans (Spruijt et al. 2006; Gorman et al. 2016).

LHON is characterized by degeneration of retinal ganglion cells and optic nerve atrophy, leading to vision loss (Yu-Wai-Man et al. 2011). More than half of cases in Europe are associated with a pathogenic variant in the mitochondrial gene *MT-ND4* that encodes subunit IV of the OXPHOS complex I, namely m.11778G>A (Wallace et al. 1988; Newman et al. 1991; Kirches 2011). Together with m.3460G>A in *MT-ND1* and m.14484T>C in *MT-ND6*, these three mtDNA point mutations account for the vast majority of LHON cases worldwide. However, pathogenic variants in nuclear genes, such as c.152A>G (p.Tyr51Cys) in the *DNAJC30*, have also recently been described as a novel, autosomal recessive cause of LHON in Central and Eastern European populations (Kieninger et al. 2022; Stenton et al. 2022; Skorczyk-Werner et al. 2023). LHON symptoms typically manifest in about half of male carriers and 10-20% of female carriers (Meyerson et al. 2015), indicating a sex bias and incomplete penetrance. Furthermore, the male-to-female ratio of symptomatic individuals varies across populations and depending on the LHON mtDNA mutation. A study of over 1,500 LHON patients, primarily from the United States, United Kingdom, Australia, and Canada, revealed that women have approximately a threefold lower risk of developing LHON compared to men and the distribution of mutations did not differ between the sexes (Poincenot et al. 2020), with earlier data from North East of England and Finish populations suggesting more prominent sex bias for the m.11778G>A variant compared to the m.3460G>A (approximately four-to fivefold vs twofold difference, respectively) (Yu-Wai-Man et al. 2003; Puomila et al. 2007). In Japan, the overall ratio has been reported to be as high as eightfold (Takano et al. 2022). The incomplete penetrance of LHON suggests that there must be additional modifying risk factors besides the primary genetic variants, which are necessary but not sufficient for LHON development.

Risk factors for LHON are usually classified as environmental (such as heavy alcohol consumption and smoking (Cullom et al. 1993; Riordan-Eva et al. 1995; Sadun et al. 2003; Kirkman et al. 2009; Hedström et al. 2024)), hormonal (e.g. the protective effect of estrogens in females (Giordano et al. 2011; Pisano et al. 2015)), and genetic. As for the latter, certain mtDNA haplogroups (i.e. sets of specific variants) are known to affect LHON penetrance, although their effect may vary by region. For example, haplogroup J is a known risk factor for LHON in some populations (Brown et al. 1997; Howell et al. 2003; Yen et al. 2006), but not in the Polish one, where haplogroup K has been proposed as a possible modifier of m.11778G>A-related disease risk (Piotrowska-Nowak et al. 2020). Western and Northern Europeans who retained vision despite carrying a pathogenic LHON mtDNA variant were found to have 30-40% more mtDNA copies in their peripheral blood than those who experienced vision loss, suggesting a lower mtDNA copy number (mtCN) as a risk factor (Giordano et al. 2014; Bianco et al. 2017, 2018). A synergistic effect between the mitochondrial m.11778G>A and the nuclear c.157C>T (p.Arg53Trp) variant in the *PRICKLE3* gene has been reported in Chinese LHON patients, in whom an earlier onset and increased penetrance of the disease have been observed (Yu et al. 2020).

The aforementioned genetic risk factors clearly demonstrate that LHON development may vary significantly by geographic region. While these associations have been described in several European populations, the high variability (Cha et al. 2015; Kowalczyk et al. 2021; Menger et al. 2021) and maternal inheritance of mtDNA warrant caution when extrapolating such findings. To date, no study has investigated the potential association between mtCN or the *PRICKLE3* c.157C>T variant and LHON penetrance in Polish individuals. The aim of this study was to perform a focused, population-specific evaluation of two previously proposed genetic modifiers of LHON penetrance. We used qPCR to examine the variability of mtCN in Polish carriers of the m.11778G>A variant to evaluate its role in LHON onset. Additionally, we screened for the presence of the c.157C>T variant in the *PRICKLE3* gene using the PCR-RFLP assay. We found that none of these proposed risk factors is associated with LHON development in the Polish population under conditions tested.

## 2 Materials and Methods

### 2.1 Study population

We analyzed a cohort of 262 individuals of Polish descent to investigate mtDNA copy number and the presence of the *PRICKLE3* c.157C>T variant. DNA samples from patients and their relatives were referred from the Medical University of Warsaw and the Center for Medical Genetics GENESIS. In all cases, total DNA extracted from peripheral blood was used. The study population was divided into three groups.

The first study group consisted of 65 men and 4 women (*N*_1_ = 69) who experienced vision loss and were molecularly confirmed to carry the pathogenic mtDNA variant m.11778G>A, associated with LHON. The average age in this group was 29.3 ± 10.4 years.

The second study group included 6 men and 25 women (*N*_2_ = 31) who, at the time of genetic testing, carried the m.11778G>A variant but showed no symptoms of LHON. Most were relatives of patients from the first study group. The average age in this group was 39.7 ± 16.1 years. Penetrance was calculated as the proportion of symptomatic individuals among all carriers of the m.11778G>A variant, stratified by sex.

For mtCN analysis, we used a control group of 68 healthy male subjects of similar age (mean age 21.8 ± 1.1 years), described previously (Piotrowska-Nowak et al. 2023). For *PRICKLE3* variant analysis, the control group comprised 94 individuals (38 females and 56 males, mean age 71.8 ± 8.8 years) who were being treated for simple senile cataract. LHON and other intraocular pathologies were excluded (Piotrowska-Nowak et al. 2019). Pathogenic mtDNA changes were excluded using next-generation sequencing, as described in detail previously (Piotrowska-Nowak et al. 2019, 2023). Control subjects in both groups were unrelated.

### 2.2 Determination of mtDNA copy number using qPCR

Quantification of absolute mtCN was performed using real-time PCR on a LightCycler 480 system with SYBR Green I chemistry (Roche), following the protocol previously described by Piotrowska-Nowak et al. (Piotrowska-Nowak et al. 2023). Briefly, the mitochondrial *MT-ND1* gene and the nuclear *B2M* gene were amplified in parallel, and mtCN was calculated as the ratio of *MT-ND1* to *B2M* gene copies, as normalized to the haploid nuclear genome. Each reaction was run in triplicate using two DNA inputs (2 ng and 0.2 ng) to ensure quantification within the linear detection range. Standard curves generated from plasmid DNA containing single-copy inserts of the target genes were included on each plate, and a synthetic oligonucleotide calibrator containing equimolar amounts of *MT-ND1* and *B2M* fragments was used to control for inter– and intra-assay variability.

### 2.3 Detection of the PRICKLE3 c.157C>T variant using PCR-RFLP

To investigate the presence of the *PRICKLE3* gene variant c.157C>T, a PCR reaction was performed using the following primers: forward 5^′^-CTTCGTCTCTAACTCTGAGCGC and reverse 5^′^-CAGAAATAGGTTCCCACCTGAG (GenBank reference sequence NG_017135.2). Reactions were performed in 25 µl volume using PCR Mix (A&A Biotechnology) following the manufacturer’s procedure with a final primer concentration of 2 µM. The thermal cycling conditions were as follows: initial denaturation at 94 ^◦^C for 4 min; 35 cycles of 94 °C for 30 s, 60 °C for 30 s, and 72 °C for 45 s; and a final extension at 72 °C for 5 min. To detect the c.157C>T variant, restriction fragment length polymorphism (RFLP) analysis was performed. This substitution abolishes the recognition site for the *Sac*II restriction enzyme. A volume of 1 µl of *Sac*II enzyme (20 U/µl, New England Biolabs) was added directly to 10 µl of the PCR product, and the mixture was incubated overnight at 37 ^◦^C. Digestion products were separated on a 1% agarose gel containing ethidium bromide and visualized under UV light. In the presence of the c.157C>T variant, the uncut PCR product is 724 bp long. In the wild-type sequence, the restriction enzyme cleaves the PCR product into two fragments of 475 bp and 249 bp.

### 2.4 Statistical analysis

Mean mtCN and standard deviation (SD) were calculated for each study group. The Shapiro-Wilk test was used to assess the normality of data distribution within each group. As the assumptions of normality were not met, differences in mtCN distribution between symptomatic carriers, asymptomatic carriers, and control individuals, as well as between males and females were evaluated using the non-parametric Kruskal-Wallis test and Mann-Whitney U test, respectively. A significance threshold of *p* < 0.05 was applied each time, unless specified otherwise. To assess the homogeneity of variances of mtCN among the three groups, the Brown-Forsythe test and subsequent pairwise comparisons with the Bonferroni correction were performed. An adjusted significance threshold of *p* < 0.0167 was applied. Spearman’s rank correlation tests were performed to explore a possible link between age and mtCN in all subjects together and separately in all study groups. An adjusted significance threshold of *p* < 0.0125 was applied. For symptomatic carriers, the age at the time of medical referral was used as the approximate age of LHON symptom onset (Table S1). Stratified analyses (e.g. by sex, mitochondrial haplogroup background, or environmental factors) were not performed due to limited subgroup sizes, which would have resulted in insufficient statistical power.

## 3 Results

In our cohort, the penetrance of LHON symptoms among Polish carriers of the m.11778G>A variant was estimated at 91.5% in males and 13.8% in females, corresponding to a 6.6-fold higher risk of disease manifestation in males. To further understand potential contributing factors, we determined the absolute mtCN in peripheral blood by calculating the ratio between the mitochondrial *MT-ND1* gene and the nuclear *B2M* gene, using copy number reference standards. The quantified mtCN values ranged from 24 to 207 in symptomatic carriers, from 33 to 138 in asymptomatic carriers, and from 52 to 123 in the control group. As shown in Table 1, the mean mtCN was 88 ± 39.51 (median: 77) in symptomatic carriers of the m.11778G>A, 75 ± 20.17 (median: 73) in asymptomatic carriers, and 82 ± 17.01 (median: 80) in healthy controls. The Shapiro-Wilk test indicated that mtCN was not normally distributed within symptomatic carriers (*W* = 0.93, *p* = 1.1 × 10^−3^), asymptomatic carriers (*W* = 0.89, *p* = 4.0 × 10^−3^) and the control group (*W* = 0.96, *p* = 0.036). The Kruskal-Wallis test revealed no statistically significant differences in mtCN among the three groups (*H* = 3.28, *p* = 0.19) (Fig. 1). Nor were any significant differences observed between men and women (*U* = 1529.5, *p* = 0.13). Individual mtCN values are presented in Online Resource Table S1.

**Fig. 1.**
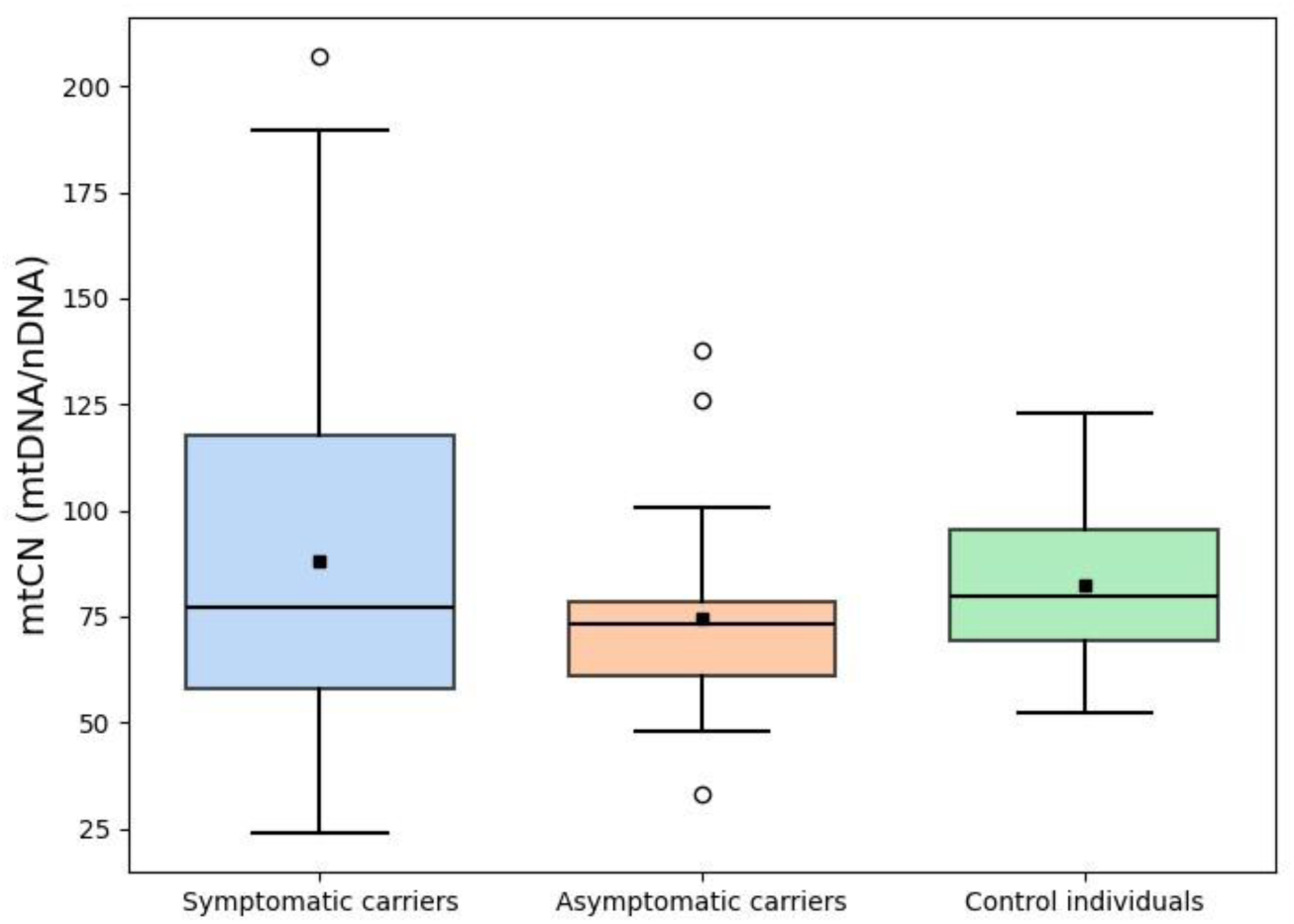
Box plots showing mtCN in peripheral blood of symptomatic carriers of the m.11778G>A variant, asymptomatic carriers, and healthy controls. Comparison of mtCN among the groups was performed using Kruskal-Wallis test and no statistically significant differences were found (*H* = 3.28, *p* = 0.194)

**Table 1.** Variability of mtCN in peripheral blood across three study groups: symptomatic carriers of the m.11778G>A variant, asymptomatic carriers, and healthy individuals without pathogenic changes in mtDNA. Results are presented separately for males (M) and females (F), as well as combined (M and F) if applicable

| Group | Sex | N | Mean mtCN ± SD | mtCN median |
| --- | --- | --- | --- | --- |
| Symptomatic carriers | M | 65 | 87 ± 40.01 | 77 |
| | F | 4 | 99 ± 27.93 | 93 |
| | M and F | 69 | 88 ± 39.51 | 77 |
| Asymptomatic carriers | M | 6 | 83 ± 12.49 | 76 |
| | F | 25 | 73 ± 21.18 | 70 |
| | M and F | 31 | 75 ± 20.17 | 73 |
| Control individuals | M | 68 | 82 ± 17.01 | 80 |

Differences in variance of mtCN between the three groups were assessed using the Brown-Forsythe test, which indicated significant heterogeneity of variances (*F* = 14.91, *p* = 1.1 × 10^−6^). Pairwise comparisons revealed significantly greater variance in the symptomatic carriers (*s*^2^ = 1584.28) compared to both the control group (*s*^2^ = 293.77; *F* = 10.98, *p* = 1.3 × 10^−3^) and the asymptomatic carriers (*s*^2^ = 420.46; *F* = 22.34, *p* = 5.6×10^−6^). No significant difference in variance was observed between the asymptomatic carriers and control individuals (*F* = 0.12, *p* = 0.74).

Spearman’s rank correlation tests were performed to evaluate the relationship between age and mtCN. Among symptomatic carriers, no statistically significant association was observed between mtCN and the approximate age of symptom onset (ρ = 0.114, *p* = 0.371; Online Resource Fig. S1). Similarly, no significant correlation was found between age and mtCN in asymptomatic carriers (ρ = 0.084, *p* = 0.690), control individuals (ρ = 0.031, *p* = 0.802), or the entire cohort (ρ = 0.004, *p* = 0.957), suggesting that age was not significantly associated with mtCN levels in this dataset.

In addition, we designed and applied a PCR-RFLP assay to detect the *PRICKLE3* c.157C>T variant, previously reported as a potential modifier of LHON penetrance (Yu et al. 2020). In this study, the c.157C>T variant was not detected in any of the 194 tested samples: 69 LHON patients carrying m.11778G>A, 31 asymptomatic carriers, and 94 control individuals. Given that *PRICKLE3* is located on the X chromosome, and that the cohort included 67 females (two alleles each) and 127 males (one allele each), a total of 261 X chromosomes were analyzed. Based on this number, it can be concluded with 95% confidence that the allele frequency is less than 1.15%, indicating that this variant is relatively rare in the Polish population.

## 4 Discussion

Mitochondrial dysfunction is implicated in a broad spectrum of human diseases, from neurodegenerative disorders and metabolic syndromes to cancer and aging. Mutations in the mitochondrial genome can severely impair mitochondrial function and contribute to both inherited and sporadic diseases. Leber hereditary optic neuropathy, a genetic disorder characterized by acute or subacute vision loss, exemplifies a condition caused primarily by pathogenic mtDNA variants. However, its incomplete penetrance and strong sex bias strongly suggest the involvement of additional genetic and environmental modifying factors. This study aimed to investigate two such potential modifiers – mtDNA copy number and the *PRICKLE3* gene variant c.157C>T (p.Arg53Trp) in a Polish cohort.

We quantified mtCN in peripheral blood as a practical alternative to tissue unavailable for testing (i.e. optic nerve) in Polish individuals carrying the m.11778G>A mutation. Importantly, blood-based mtCN assessment was used previously to test a proposed risk factor hypothesis in a comparable manner. Previous research in Spanish, Italian, and British populations suggested a protective effect of higher mtCN in LHON: unaffected carriers were reported to have 30-50% more mtDNA copies than symptomatic individuals (Giordano et al. 2014; Bianco et al. 2018). To our knowledge, such an analysis had not previously been performed in a Polish LHON cohort. However, our findings do not support this association; we report that mtCN does not significantly influence the manifestation of LHON symptoms in Polish m.11778G>A carriers under the conditions tested.

While our analysis showed no significant differences in the central tendency (mean or median) of mtCN between the groups, the significantly higher variance observed in symptomatic carriers is particularly noteworthy and suggests that mtCN responses may not be uniform within this group. This broader and more diverse distribution of mtCN values indicates substantial inter-individual heterogeneity in affected subjects. One possible explanation is that, in response to mitochondrial dysfunction, symptomatic individuals may activate compensatory mechanisms such as increased mitochondrial biogenesis (Moreno-Loshuertos et al. 2006; Korsten et al. 2010), which could lead to elevated mtCN. However, the effectiveness of this homeostatic response may vary substantially between individuals, resulting in a wide range of mtCN values observed among symptomatic carriers and potentially masking an underlying association between mtCN levels and LHON expression when standard comparisons are applied.

Age is another important confounding factor. LHON manifests most often in early adulthood (Poincenot et al. 2020), while mtCN tends to decline with age (Wachsmuth et al. 2016; Zhang et al. 2017; Liu et al. 2021). Our asymptomatic group was, on average, a decade older than the symptomatic group, which may have masked a potential protective effect of higher mtCN. However, this may be more subtle or negligible, as previous studies have shown that individuals between the ages of 18 and 48 years have similar mtCN (Mengel-From et al. 2014). Consistent with this, our correlation analyzes did not reveal a significant relationship between mtCN and age – neither when approximating the age of LHON onset in symptomatic individuals nor when examining age in any other of the study groups – supporting the conclusion that age-related differences likely did not influence mtCN results in our study.

We also employed a PCR-RFLP assay to screen for the *PRICKLE3* c.157C>T variant, which has been previously implicated in modulating LHON penetrance. In the Chinese population, this variant showed a synergistic effect with the *MT-ND4* m.11778G>A mutation, exacerbating OXPHOS dysfunction (Yu et al. 2020). However, consistent with the findings in unspecified Caucasian populations (Yu et al. 2020), we did not detect the c.157C>T variant in any of the Polish individuals analyzed. These results suggest that this variant is unlikely to represent a major modifier of LHON penetrance in the Polish population.

According to the gnomAD data, the reported allele frequency of c.157C>T is 2.2 × 10^−6^ in Europe and 3.0 × 10^−5^ in East Asia (Karczewski et al. 2020). Such interpopulation variant frequency differences may explain its absence in our cohort and highlight the importance of population-specific genetic analyses. Evolutionary mechanisms, including natural selection, genetic drift, and historical migration patterns (Ingram et al. 2009; Vuorisalo et al. 2012), may underlie this variability.

Mitochondrial haplogroup background is another potential source of population-specific findings. For the m.11778G>A variant, increased LHON risk has been associated with haplogroup J2, while haplogroup H confers a protective effect (Brown et al. 1997). Both haplogroups are common in Europe, including Poland (Lott et al. 2013; Jarczak et al. 2019; Kaja et al. 2022). Our group has previously reported no association between haplogroup J and m.11778G>A penetrance in Polish LHON patients (Piotrowska-Nowak et al. 2020). Haplogroup-defining variants may also affect cellular mtCN: cybrid studies have shown that haplogroup J1b is associated with higher mtCN than haplogroup H, while J1c and J2 are linked to lower mtCN (Suissa et al. 2009; Gómez-Durán et al. 2010). Although in the present study haplogroup stratification was not performed, major differences in the distribution of mtCN-inluencing haplogroups between the study groups are unlikely, as most asymptomatic carriers were maternally related to symptomatic individuals and therefore shared the same haplogroup. Nevertheless, we cannot fully exclude that subtle haplogroup-related effects may have contributed to inter-individual variability in mtCN and potentially obscured weak associations between mtCN and LHON symptom manifestation, specifically a protective effect of higher mtCN, particularly given the limited sample size.

Moreover, the haplogroup distribution in Poland differs markedly from that in East Asia, including China, where D and M haplogroups are dominant (Rishishwar and Jordan 2017). In particular, haplogroup M7b has been associated with an increased risk of vision loss, while haplogroups M8a and F have protection against LHON among carriers of m.11778G>A (Ji et al. 2008; Zhang et al. 2011). While nuclear–mitochondrial interactions have been observed in genome-wide studies (e.g. within-haplogroup GWAS) (Xia et al. 2023), specific link between nuclear variants like *PRICKLE3* c.157C>T and mtDNA haplogroups has not been demonstrated. However, if such linkage is present with a haplogroup that is rare or absent in the Polish population, it would further explain variant absence in our cohort.

This study has several limitations. First, the sample size – 168 individuals for mtCN analysis and 194 for *PRICKLE3* screening – was relatively modest, though comparable to previous LHON studies (Giordano et al. 2014; Bianco et al. 2018). This is a common challenge, given the low prevalence of LHON (Yu-Wai-Man et al. 2003). Second, in our cohort, the prevalence of disease manifestation among m.11778G>A carriers was approximately 7-fold higher in males. Although this male-to-female ratio somewhat exceeds the typically reported 3– to 5-fold difference observed in European LHON cohorts (Meyerson et al. 2015), such variability is not unexpected and may result from population-specific factors as well as differences in study design, including limited sample size. The high penetrance observed in males in our cohort (91.5%) may also reflect a bias in estimation, as symptomatic individuals are more likely to be referred for genetic testing, while asymptomatic male carriers may remain undiagnosed. This could lead to an overestimation of penetrance in men. However, due to the substantial disparity in the number of male and female participants within each study group, a detailed mtCN analysis by sex was not feasible in this study, as it would lack sufficient statistical power. Finally, environmental factors such as smoking, which has been shown to reduce mtCN in both LHON carriers and healthy individuals (Kirkman et al. 2009; Vyas et al. 2020), were not controlled for as such information was not systematically collected, and we cannot exclude their influence on the observed results.

Taken together, our findings suggest that neither the *PRICKLE3* c.157C>T variant nor mtDNA copy number substantially influences LHON manifestation in the Polish population under conditions tested. These results imply that the genetic basis of LHON and its modifying factors are more complex and geographically diverse than initially thought. Geographic variation in genetic disease expression is not unique to LHON. Notable examples include sickle cell anemia (Piel et al. 2013) and G6PD deficiency (Howes et al. 2012; Võ et al. 2024), both prevalent in malaria-endemic regions; Tay-Sachs disease, more common in specific founder populations such as Ashkenazi Jews (Rozenberg and Pereira 2001); and cystic fibrosis, which shows marked regional heterogeneity in *CFTR* variant distribution across Russia (Petrova et al. 2021).

The molecular mechanisms underlying the variable penetrance of LHON-associated mtDNA mutations remain largely unresolved. LHON is likely a multifactorial disease in which symptom expression depends on the interplay of several genetic and environmental factors. The results of this study highlight the need for large-scale, population-specific studies that consider these multiple contributing factors. Future research in Polish and other populations should aim to simultaneously assess mtDNA copy number, haplogroup background, nuclear modifiers, and environmental exposures to gain a more comprehensive understanding of the etiology of LHON.

## Statements & Declarations

### Funding

This research did not receive any specific grant from funding agencies in the public, commercial, or not-for-profit sectors. It was supported by the statutory institutional funding of the University of Warsaw.

### Ethics Approval

The study was approved by the Ethics Committee of the Medical University of Warsaw, Poland (approval number KB/187/2015) and was performed in accordance with the ethical standards of the Declaration of Helsinki 1964 and its later amendments. Informed consent was obtained from all individuals included in the study.

### Competing Interests

none.

## Author Contributions

Julia Sikorska: Methodology, Investigation, Formal analysis, Writing – Original Draft.

Maciej R. Krawczyński: Resources.

Magdalena Korwin: Resources.

Monika Ołdak: Resources, Writing – Review & Editing.

Ewa Bartnik: Supervision, Writing – Review & Editing.

Katarzyna Tońska: Supervision, Writing – Review & Editing.

Agnieszka Piotrowska-Nowak: Methodology, Conceptualization, Validation, Writing – Review & Editing, Supervision.

Each author made a substantial contribution to the work, drafted or critically revised the manuscript, and approved the final version.

## Data Availability

The data generated and/or analyzed during the current study are included in this published article and its supplementary information files, or are available from the corresponding author on reasonable request.

## Notes

### Competing Interest Statement

The authors have declared no competing interest.

### Author Declarations

The Ethics Committee of the Medical University of Warsaw, Poland, gave ethical approval for this work (number KB/187/2015).

