## Supplementary material for "Analysis of genetic risk factors for Leber hereditary optic neuropathy in the Polish population"

### Online Resource

Julia Sikorska[a,1], Maciej R. Krawczyński[b,c], Magdalena Korwin[d], Monika Ołdak[e,f], Ewa Bartnik[a], Katarzyna Tońska[a], Agnieszka Piotrowska-Nowak [a*]

[a] Institute of Genetics and Biotechnology, Faculty of Biology, University of Warsaw, 02-106 Warsaw, Poland

 (JS), (EB), (KT), (APN)

[b] Department of Medical Genetics, Poznan University of Medical Sciences, 60-806 Poznan, Poland

 (MRK)

[c] Center for Medical Genetics GENESIS, 60-529 Poznan, Poland

[d] Department of Ophthalmology, Medical University of Warsaw, 02-005 Warsaw, Poland

 (MK)

[e] Department of Genetics, Institute of Physiology and Pathology of Hearing, 02-042 Warsaw, Poland

 (MO)

[f] Department of Histology and Embryology, Medical University of Warsaw, 02-004 Warsaw, Poland

[1] Present address: Interdisciplinary Laboratory of Biological Systems Modelling, Centre of New Technologies, University of Warsaw, Banacha 2c, 02-097 Warsaw, Poland

* Correspondence:

Agnieszka Piotrowska-Nowak

University of Warsaw, Faculty of Biology, Institute of Genetics and Biotechnology

Pawińskiego 5a Street, 02-106 Warsaw, Poland

+48 22 592 22 39

### List of Tables

S1 Demographic and molecular data for each individual included in the study. . . . . . . . . . . . . . . . . . . . . . . . . . S1

### List of Figures

S1 A scatter plot illustrating the relationship between the approximate age of LHON symptom onset and mtCN in symptomatic carriers. Spearman's rank correlation test revealed a correlation coefficient ρ of 0.114 with a *p*-value of 0.371, indicating no statistically significant association. . . . . . . . . . . . . . . . . . . . . . . . . . . . . . . S14

**Table S1** Demographic and molecular data for each individual included in the study

| Study subject | Group | Tissue | Age (year range)[a] | Sex | mtDNA copy number (mean *MT-ND1*/*B2M*) | mtDNA copy number (SD) | Presence of *PRICKLE3* variant (c.157C>A) (yes/no) |
|---|---|---|---|---|---|---|---|
| 1 | Symptomatic carriers | blood | 26-30 | M | 189.85 | 27.74 | no |
| 2 | Symptomatic carriers | blood | 16-20 | M | 104.32 | 14.66 | no |
| 3 | Symptomatic carriers | blood | n/a | M | 48.97 | 8.90 | no |
| 4 | Symptomatic carriers | blood | 26-30 | M | 148.03 | 13.46 | no |
| 5 | Symptomatic carriers | blood | 6-10 | M | 63.48 | 13.65 | no |
| 6 | Symptomatic carriers | blood | 31-35 | M | 135.00 | 4.32 | no |
| 7 | Symptomatic carriers | blood | 26-30 | M | 88.14 | 9.75 | no |
| 8 | Symptomatic carriers | blood | 41-45 | M | 65.04 | 12.61 | no |
| 9 | Symptomatic carriers | blood | 21-25 | M | 207.07 | 27.76 | no |
| 10 | Symptomatic carriers | blood | 21-25 | M | 136.13 | 7.38 | no |
| 11 | Symptomatic carriers | blood | 21-25 | M | 49.62 | 0.02 | no |
| 12 | Symptomatic carriers | blood | 16-20 | M | 49.83 | 3.21 | no |
| 13 | Symptomatic carriers | blood | 41-45 | M | 71.43 | 1.26 | no |
| 14 | Symptomatic carriers | blood | 21-25 | M | 80.66 | 22.42 | no |
| 15 | Symptomatic carriers | blood | 16-20 | M | 82.97 | 10.39 | no |
| 16 | Symptomatic carriers | blood | 31-35 | M | 50.92 | 12.10 | no |
| 17 | Symptomatic carriers | blood | 21-25 | M | 71.59 | 4.12 | no |

| | | | | | | | |
|---|---|---|---|---|---|---|---|
| 18 | Symptomatic carriers | blood | 11-15 | M | 59.50 | 7.51 | no |
| 19 | Symptomatic carriers | blood | 31-35 | M | 44.69 | 1.64 | no |
| 20 | Symptomatic carriers | blood | 31-35 | M | 77.03 | 7.62 | no |
| 21 | Symptomatic carriers | blood | 36-40 | M | 115.93 | 10.93 | no |
| 22 | Symptomatic carriers | blood | 21-25 | M | 165.11 | 11.06 | no |
| 23 | Symptomatic carriers | blood | 21-25 | M | 106.55 | 1.05 | no |
| 24 | Symptomatic carriers | blood | 21-25 | M | 39.10 | 0.67 | no |
| 25 | Symptomatic carriers | blood | 31-35 | M | 69.00 | 2.15 | no |
| 26 | Symptomatic carriers | blood | 16-20 | M | 77.03 | 1.22 | no |
| 27 | Symptomatic carriers | blood | 16-20 | M | 150.55 | 4.37 | no |
| 28 | Symptomatic carriers | blood | 36-40 | M | 120.18 | 16.08 | no |
| 29 | Symptomatic carriers | blood | 21-25 | M | 60.92 | 5.66 | no |
| 30 | Symptomatic carriers | blood | n/a | M | 37.07 | 2.55 | no |
| 31 | Symptomatic carriers | blood | 26-30 | M | 57.51 | 0.72 | no |
| 32 | Symptomatic carriers | blood | 31-35 | M | 46.37 | 1.45 | no |
| 33 | Symptomatic carriers | blood | 11-15 | M | 30.61 | 3.98 | no |
| 34 | Symptomatic carriers | blood | 21-25 | M | 24.00 | 0.30 | no |
| 35 | Symptomatic carriers | blood | 16-20 | M | 52.58 | 11.97 | no |
| 36 | Symptomatic carriers | blood | 26-30 | M | 74.60 | 3.23 | no |
| 37 | Symptomatic carriers | blood | 41-45 | M | 59.35 | 4.29 | no |

| | | | | | | | |
|---|---|---|---|---|---|---|---|
| 38 | Symptomatic carriers | blood | 31-35 | M | 142.93 | 1.24 | no |
| 39 | Symptomatic carriers | blood | 31-35 | M | 63.97 | 13.40 | no |
| 40 | Symptomatic carriers | blood | 31-35 | M | 117.63 | 0.67 | no |
| 41 | Symptomatic carriers | blood | 31-35 | M | 85.33 | 18.08 | no |
| 42 | Symptomatic carriers | blood | n/a | M | 77.45 | 0.26 | no |
| 43 | Symptomatic carriers | blood | 31-35 | M | 49.29 | 4.90 | no |
| 44 | Symptomatic carriers | blood | 51-55 | M | 159.44 | 9.31 | no |
| 45 | Symptomatic carriers | blood | 46-50 | M | 65.83 | 6.65 | no |
| 46 | Symptomatic carriers | blood | 26-30 | M | 125.81 | 14.34 | no |
| 47 | Symptomatic carriers | blood | 16-20 | M | 46.69 | 2.07 | no |
| 48 | Symptomatic carriers | blood | 21-25 | M | 122.52 | 7.95 | no |
| 49 | Symptomatic carriers | blood | 16-20 | M | 122.35 | 10.59 | no |
| 50 | Symptomatic carriers | blood | n/a | M | 82.30 | 3.19 | no |
| 51 | Symptomatic carriers | blood | 36-40 | M | 56.08 | 2.54 | no |
| 52 | Symptomatic carriers | blood | 26-30 | M | 90.66 | 18.28 | no |
| 53 | Symptomatic carriers | blood | 16-20 | M | 109.48 | 32.35 | no |
| 54 | Symptomatic carriers | blood | 26-30 | M | 131.21 | 29.88 | no |
| 55 | Symptomatic carriers | blood | 11-15 | M | 48.87 | 5.84 | no |
| 56 | Symptomatic carriers | blood | 31-35 | M | 68.55 | 11.44 | no |
| 57 | Symptomatic carriers | blood | 36-40 | M | 101.10 | 1.95 | no |

| | | | | | | | |
|---|---|---|---|---|---|---|---|
| 58 | Symptomatic carriers | blood | 36-40 | M | 141.57 | 7.53 | no |
| 59 | Symptomatic carriers | blood | 56-60 | M | 134.49 | 12.77 | no |
| 60 | Symptomatic carriers | blood | 16-20 | M | 67.41 | 5.37 | no |
| 61 | Symptomatic carriers | blood | 26-30 | M | 86.00 | 13.17 | no |
| 62 | Symptomatic carriers | blood | 31-35 | M | 56.67 | 10.04 | no |
| 63 | Symptomatic carriers | blood | 31-35 | M | 76.91 | 8.00 | no |
| 64 | Symptomatic carriers | blood | 41-45 | M | 77.55 | 20.07 | no |
| 65 | Symptomatic carriers | blood | 26-30 | M | 58.20 | 3.93 | no |
| 66 | Symptomatic carriers | blood | 31-35 | F | 141.93 | 6.44 | no |
| 67 | Symptomatic carriers | blood | n/a | F | 106.31 | 0.77 | no |
| 68 | Symptomatic carriers | blood | 46-50 | F | 80.43 | 4.78 | no |
| 69 | Symptomatic carriers | blood | 51-55 | F | 69.32 | 0.09 | no |
| 70 | Asymptomatic carriers | blood | 16-20 | M | 74.72 | 3.96 | no |
| 71 | Asymptomatic carriers | blood | 21-25 | M | 100.69 | 7.15 | no |
| 72 | Asymptomatic carriers | blood | 11-15 | M | 73.96 | 6.86 | no |
| 73 | Asymptomatic carriers | blood | n/a | M | 99.44 | 23.47 | no |
| 74 | Asymptomatic carriers | blood | 26-30 | M | 76.68 | 6.17 | no |
| 75 | Asymptomatic carriers | blood | 41-45 | M | 70.21 | 0.04 | no |
| 76 | Asymptomatic carriers | blood | 41-45 | F | 73.23 | 4.90 | no |
| 77 | Asymptomatic carriers | blood | 26-30 | F | 58.47 | 2.08 | no |

| | | | | | | | |
|---|---|---|---|---|---|---|---|
| 78 | Asymptomatic carriers | blood | 66-70 | F | 78.19 | 9.97 | no |
| 79 | Asymptomatic carriers | blood | 41-45 | F | 75.72 | 3.14 | no |
| 80 | Asymptomatic carriers | blood | 46-50 | F | 69.33 | 17.19 | no |
| 81 | Asymptomatic carriers | blood | 21-25 | F | 78.75 | 10.47 | no |
| 82 | Asymptomatic carriers | blood | 31-35 | F | 47.92 | 7.88 | no |
| 83 | Asymptomatic carriers | blood | n/a | F | 87.77 | 14.11 | no |
| 84 | Asymptomatic carriers | blood | 46-50 | F | 77.76 | 16.84 | no |
| 85 | Asymptomatic carriers | blood | n/a | F | 125.94 | 8.81 | no |
| 86 | Asymptomatic carriers | blood | 21-25 | F | 57.82 | 7.71 | no |
| 87 | Asymptomatic carriers | blood | n/a | F | 33.17 | 0.94 | no |
| 88 | Asymptomatic carriers | blood | 41-45 | F | 60.38 | 1.59 | no |
| 89 | Asymptomatic carriers | blood | 56-60 | F | 60.86 | 2.78 | no |
| 90 | Asymptomatic carriers | blood | 36-40 | F | 71.04 | 8.05 | no |
| 91 | Asymptomatic carriers | blood | 21-25 | F | 59.34 | 5.89 | no |
| 92 | Asymptomatic carriers | blood | 21-25 | F | 61.27 | 6.28 | no |
| 93 | Asymptomatic carriers | blood | 61-65 | F | 69.51 | 8.63 | no |
| 94 | Asymptomatic carriers | blood | n/a | F | 77.86 | 4.05 | no |
| 95 | Asymptomatic carriers | blood | 36-40 | F | 65.36 | 7.14 | no |
| 96 | Asymptomatic carriers | blood | 36-40 | F | 137.84 | 31.60 | no |
| 97 | Asymptomatic carriers | blood | 61-65 | F | 85.39 | 1.22 | no |

| | | | | | | | |
|---|---|---|---|---|---|---|---|
| 98 | Asymptomatic carriers | blood | n/a | F | 63.97 | 7.15 | no |
| 99 | Asymptomatic carriers | blood | 66-70 | F | 60.06 | 8.42 | no |
| 100 | Asymptomatic carriers | blood | 56-60 | F | 82.71 | 1.42 | no |
| 101 | Control group for mtCN | blood | 21-25 | M | 63.98 | 1.42 | n/a |
| 102 | Control group for mtCN | blood | 21-25 | M | 65.58 | 9.98 | n/a |
| 103 | Control group for mtCN | blood | 21-25 | M | 93.94 | 14.21 | n/a |
| 104 | Control group for mtCN | blood | 21-25 | M | 69.68 | 4.26 | n/a |
| 105 | Control group for mtCN | blood | 21-25 | M | 70.47 | 7.51 | n/a |
| 106 | Control group for mtCN | blood | 21-25 | M | 53.01 | 8.49 | n/a |
| 107 | Control group for mtCN | blood | 21-25 | M | 52.29 | 5.79 | n/a |
| 108 | Control group for mtCN | blood | 21-25 | M | 67.64 | 2.37 | n/a |
| 109 | Control group for mtCN | blood | 21-25 | M | 96.78 | 0.00 | n/a |
| 110 | Control group for mtCN | blood | 21-25 | M | 70.83 | 12.56 | n/a |
| 111 | Control group for mtCN | blood | 21-25 | M | 66.02 | 0.75 | n/a |
| 112 | Control group for mtCN | blood | 21-25 | M | 67.31 | 7.45 | n/a |
| 113 | Control group for mtCN | blood | 21-25 | M | 77.03 | 7.17 | n/a |
| 114 | Control group for mtCN | blood | 21-25 | M | 93.64 | 3.19 | n/a |
| 115 | Control group for mtCN | blood | 21-25 | M | 69.29 | 5.02 | n/a |
| 116 | Control group for mtCN | blood | 21-25 | M | 58.93 | 4.06 | n/a |
| 117 | Control group for mtCN | blood | 21-25 | M | 83.49 | 3.52 | n/a |

| | | | | | | | |
|---|---|---|---|---|---|---|---|
| 118 | Control group for mtCN | blood | 21-25 | M | 100.74 | 20.21 | n/a |
| 119 | Control group for mtCN | blood | 21-25 | M | 95.89 | 18.53 | n/a |
| 120 | Control group for mtCN | blood | 21-25 | M | 87.28 | 1.02 | n/a |
| 121 | Control group for mtCN | blood | 21-25 | M | 123.01 | 6.57 | n/a |
| 122 | Control group for mtCN | blood | 21-25 | M | 63.28 | 6.79 | n/a |
| 123 | Control group for mtCN | blood | 21-25 | M | 91.46 | 24.03 | n/a |
| 124 | Control group for mtCN | blood | 21-25 | M | 107.22 | 12.09 | n/a |
| 125 | Control group for mtCN | blood | 21-25 | M | 103.90 | 24.69 | n/a |
| 126 | Control group for mtCN | blood | 21-25 | M | 73.33 | 0.87 | n/a |
| 127 | Control group for mtCN | blood | 21-25 | M | 95.26 | 11.26 | n/a |
| 128 | Control group for mtCN | blood | 21-25 | M | 97.60 | 12.15 | n/a |
| 129 | Control group for mtCN | blood | 21-25 | M | 83.06 | 5.46 | n/a |
| 130 | Control group for mtCN | blood | 21-25 | M | 92.79 | 9.88 | n/a |
| 131 | Control group for mtCN | blood | 21-25 | M | 91.73 | 13.52 | n/a |
| 132 | Control group for mtCN | blood | 21-25 | M | 109.30 | 4.28 | n/a |
| 133 | Control group for mtCN | blood | 21-25 | M | 103.26 | 10.27 | n/a |
| 134 | Control group for mtCN | blood | 21-25 | M | 82.21 | 4.32 | n/a |
| 135 | Control group for mtCN | blood | 21-25 | M | 101.08 | 18.81 | n/a |
| 136 | Control group for mtCN | blood | 21-25 | M | 70.03 | 1.77 | n/a |
| 137 | Control group for mtCN | blood | 21-25 | M | 66.02 | 0.09 | n/a |

| | | | | | | | |
|---|---|---|---|---|---|---|---|
| 138 | Control group for mtCN | blood | 21-25 | M | 116.56 | 8.35 | n/a |
| 139 | Control group for mtCN | blood | 21-25 | M | 84.15 | 9.31 | n/a |
| 140 | Control group for mtCN | blood | 21-25 | M | 106.09 | 4.65 | n/a |
| 141 | Control group for mtCN | blood | 21-25 | M | 91.55 | 2.78 | n/a |
| 142 | Control group for mtCN | blood | 21-25 | M | 115.51 | 25.73 | n/a |
| 143 | Control group for mtCN | blood | 21-25 | M | 92.42 | 7.30 | n/a |
| 144 | Control group for mtCN | blood | 21-25 | M | 89.18 | 1.71 | n/a |
| 145 | Control group for mtCN | blood | 21-25 | M | 79.65 | 7.88 | n/a |
| 146 | Control group for mtCN | blood | 21-25 | M | 63.39 | 3.85 | n/a |
| 147 | Control group for mtCN | blood | 21-25 | M | 73.83 | 3.60 | n/a |
| 148 | Control group for mtCN | blood | 21-25 | M | 70.09 | 1.37 | n/a |
| 149 | Control group for mtCN | blood | 21-25 | M | 110.51 | 6.70 | n/a |
| 150 | Control group for mtCN | blood | 21-25 | M | 76.26 | 6.12 | n/a |
| 151 | Control group for mtCN | blood | 21-25 | M | 69.19 | 3.86 | n/a |
| 152 | Control group for mtCN | blood | 21-25 | M | 108.44 | 11.65 | n/a |
| 153 | Control group for mtCN | blood | 21-25 | M | 63.86 | 7.51 | n/a |
| 154 | Control group for mtCN | blood | 16-20 | M | 77.27 | 9.58 | n/a |
| 155 | Control group for mtCN | blood | 21-25 | M | 71.89 | 3.11 | n/a |
| 156 | Control group for mtCN | blood | 16-20 | M | 102.62 | 15.06 | n/a |
| 157 | Control group for mtCN | blood | 16-20 | M | 75.41 | 10.04 | n/a |

| | | | | | | | |
|---|---|---|---|---|---|---|---|
| 158 | Control group for mtCN | blood | 21-25 | M | 70.97 | 0.45 | n/a |
| 159 | Control group for mtCN | blood | 16-20 | M | 100.36 | 11.86 | n/a |
| 160 | Control group for mtCN | blood | 21-25 | M | 89.23 | 9.12 | n/a |
| 161 | Control group for mtCN | blood | 16-20 | M | 69.24 | 3.64 | n/a |
| 162 | Control group for mtCN | blood | 16-20 | M | 73.54 | 0.07 | n/a |
| 163 | Control group for mtCN | blood | 21-25 | M | 71.82 | 0.17 | n/a |
| 164 | Control group for mtCN | blood | 21-25 | M | 79.69 | 10.77 | n/a |
| 165 | Control group for mtCN | blood | 21-25 | M | 53.73 | 1.26 | n/a |
| 166 | Control group for mtCN | blood | 21-25 | M | 64.86 | 8.54 | n/a |
| 167 | Control group for mtCN | blood | 21-25 | M | 81.21 | 9.95 | n/a |
| 168 | Control group for mtCN | blood | 21-25 | M | 64.26 | 0.18 | n/a |
| 169 | Control group for *PRICKLE3* | blood | 76-80 | F | n/a | n/a | no |
| 170 | Control group for *PRICKLE3* | blood | 71-75 | F | n/a | n/a | no |
| 171 | Control group for *PRICKLE3* | blood | 71-75 | F | n/a | n/a | no |
| 172 | Control group for *PRICKLE3* | blood | 56-60 | M | n/a | n/a | no |
| 173 | Control group for *PRICKLE3* | blood | 66-70 | F | n/a | n/a | no |
| 174 | Control group for *PRICKLE3* | blood | 56-60 | M | n/a | n/a | no |
| 175 | Control group for *PRICKLE3* | blood | 81-85 | F | n/a | n/a | no |
| 176 | Control group for *PRICKLE3* | blood | 76-80 | F | n/a | n/a | no |
| 177 | Control group for *PRICKLE3* | blood | 81-85 | F | n/a | n/a | no |

| | | | | | | | |
|---|---|---|---|---|---|---|---|
| 178 | Control group for *PRICKLE3* | blood | 66-70 | F | n/a | n/a | no |
| 179 | Control group for *PRICKLE3* | blood | 61-65 | F | n/a | n/a | no |
| 180 | Control group for *PRICKLE3* | blood | 76-80 | F | n/a | n/a | no |
| 181 | Control group for *PRICKLE3* | blood | 61-65 | M | n/a | n/a | no |
| 182 | Control group for *PRICKLE3* | blood | 66-70 | F | n/a | n/a | no |
| 183 | Control group for *PRICKLE3* | blood | 71-75 | F | n/a | n/a | no |
| 184 | Control group for *PRICKLE3* | blood | 76-80 | F | n/a | n/a | no |
| 185 | Control group for *PRICKLE3* | blood | 76-80 | M | n/a | n/a | no |
| 186 | Control group for *PRICKLE3* | blood | 71-75 | F | n/a | n/a | no |
| 187 | Control group for *PRICKLE3* | blood | 56-60 | M | n/a | n/a | no |
| 188 | Control group for *PRICKLE3* | blood | 76-80 | M | n/a | n/a | no |
| 189 | Control group for *PRICKLE3* | blood | 61-65 | M | n/a | n/a | no |
| 190 | Control group for *PRICKLE3* | blood | 66-70 | F | n/a | n/a | no |
| 191 | Control group for *PRICKLE3* | blood | 56-60 | M | n/a | n/a | no |
| 192 | Control group for *PRICKLE3* | blood | 66-70 | F | n/a | n/a | no |
| 193 | Control group for *PRICKLE3* | blood | 61-65 | F | n/a | n/a | no |
| 194 | Control group for *PRICKLE3* | blood | 61-65 | F | n/a | n/a | no |
| 195 | Control group for *PRICKLE3* | blood | 81-85 | F | n/a | n/a | no |
| 196 | Control group for *PRICKLE3* | blood | 66-70 | F | n/a | n/a | no |
| 197 | Control group for *PRICKLE3* | blood | 61-65 | M | n/a | n/a | no |

| | | | | | | | |
|---|---|---|---|---|---|---|---|
| 198 | Control group for *PRICKLE3* | blood | 51-55 | M | n/a | n/a | no |
| 199 | Control group for *PRICKLE3* | blood | 66-70 | F | n/a | n/a | no |
| 200 | Control group for *PRICKLE3* | blood | 66-70 | M | n/a | n/a | no |
| 201 | Control group for *PRICKLE3* | blood | 71-75 | M | n/a | n/a | no |
| 202 | Control group for *PRICKLE3* | blood | 71-75 | M | n/a | n/a | no |
| 203 | Control group for *PRICKLE3* | blood | n/a | M | n/a | n/a | no |
| 204 | Control group for *PRICKLE3* | blood | 76-80 | F | n/a | n/a | no |
| 205 | Control group for *PRICKLE3* | blood | n/a | F | n/a | n/a | no |
| 206 | Control group for *PRICKLE3* | blood | 56-60 | F | n/a | n/a | no |
| 207 | Control group for *PRICKLE3* | blood | 76-80 | F | n/a | n/a | no |
| 208 | Control group for *PRICKLE3* | blood | 81-85 | M | n/a | n/a | no |
| 209 | Control group for *PRICKLE3* | blood | 76-80 | M | n/a | n/a | no |
| 210 | Control group for *PRICKLE3* | blood | 81-85 | M | n/a | n/a | no |
| 211 | Control group for *PRICKLE3* | blood | 66-70 | M | n/a | n/a | no |
| 212 | Control group for *PRICKLE3* | blood | 61-65 | M | n/a | n/a | no |
| 213 | Control group for *PRICKLE3* | blood | 81-85 | F | n/a | n/a | no |
| 214 | Control group for *PRICKLE3* | blood | 71-75 | F | n/a | n/a | no |
| 215 | Control group for *PRICKLE3* | blood | 76-80 | F | n/a | n/a | no |
| 216 | Control group for *PRICKLE3* | blood | 51-55 | F | n/a | n/a | no |
| 217 | Control group for *PRICKLE3* | blood | 81-85 | F | n/a | n/a | no |

| | | | | | | | |
|---|---|---|---|---|---|---|---|
| 218 | Control group for *PRICKLE3* | blood | 61-65 | F | n/a | n/a | no |
| 219 | Control group for *PRICKLE3* | blood | 66-70 | F | n/a | n/a | no |
| 220 | Control group for *PRICKLE3* | blood | 81-85 | F | n/a | n/a | no |
| 221 | Control group for *PRICKLE3* | blood | 71-75 | F | n/a | n/a | no |
| 222 | Control group for *PRICKLE3* | blood | 56-60 | F | n/a | n/a | no |
| 223 | Control group for *PRICKLE3* | blood | 61-65 | M | n/a | n/a | no |
| 224 | Control group for *PRICKLE3* | blood | 61-65 | F | n/a | n/a | no |
| 225 | Control group for *PRICKLE3* | blood | 81-85 | M | n/a | n/a | no |
| 226 | Control group for *PRICKLE3* | blood | 66-70 | F | n/a | n/a | no |
| 227 | Control group for *PRICKLE3* | blood | 76-80 | F | n/a | n/a | no |
| 228 | Control group for *PRICKLE3* | blood | 81-85 | F | n/a | n/a | no |
| 229 | Control group for *PRICKLE3* | blood | 76-80 | F | n/a | n/a | no |
| 230 | Control group for *PRICKLE3* | blood | 81-85 | M | n/a | n/a | no |
| 231 | Control group for *PRICKLE3* | blood | 81-85 | F | n/a | n/a | no |
| 232 | Control group for *PRICKLE3* | blood | 81-85 | M | n/a | n/a | no |
| 233 | Control group for *PRICKLE3* | blood | 81-85 | F | n/a | n/a | no |
| 234 | Control group for *PRICKLE3* | blood | 76-80 | M | n/a | n/a | no |
| 235 | Control group for *PRICKLE3* | blood | 86-90 | F | n/a | n/a | no |
| 236 | Control group for *PRICKLE3* | blood | 81-85 | F | n/a | n/a | no |
| 237 | Control group for *PRICKLE3* | blood | 61-65 | F | n/a | n/a | no |

| | | | | | | | |
|---|---|---|---|---|---|---|---|
| 238 | Control group for *PRICKLE3* | blood | 76-80 | F | n/a | n/a | no |
| 239 | Control group for *PRICKLE3* | blood | 76-80 | M | n/a | n/a | no |
| 240 | Control group for *PRICKLE3* | blood | 61-65 | M | n/a | n/a | no |
| 241 | Control group for *PRICKLE3* | blood | 61-65 | F | n/a | n/a | no |
| 242 | Control group for *PRICKLE3* | blood | 71-75 | M | n/a | n/a | no |
| 243 | Control group for *PRICKLE3* | blood | 66-70 | F | n/a | n/a | no |
| 244 | Control group for *PRICKLE3* | blood | 76-80 | M | n/a | n/a | no |
| 245 | Control group for *PRICKLE3* | blood | 86-90 | M | n/a | n/a | no |
| 246 | Control group for *PRICKLE3* | blood | 71-75 | F | n/a | n/a | no |
| 247 | Control group for *PRICKLE3* | blood | 66-70 | F | n/a | n/a | no |
| 248 | Control group for *PRICKLE3* | blood | 81-85 | M | n/a | n/a | no |
| 249 | Control group for *PRICKLE3* | blood | 61-65 | M | n/a | n/a | no |
| 250 | Control group for *PRICKLE3* | blood | 66-70 | M | n/a | n/a | no |
| 251 | Control group for *PRICKLE3* | blood | 61-65 | F | n/a | n/a | no |
| 252 | Control group for *PRICKLE3* | blood | 66-70 | M | n/a | n/a | no |
| 253 | Control group for *PRICKLE3* | blood | 81-85 | M | n/a | n/a | no |
| 254 | Control group for *PRICKLE3* | blood | 61-65 | F | n/a | n/a | no |
| 255 | Control group for *PRICKLE3* | blood | 66-70 | F | n/a | n/a | no |
| 256 | Control group for *PRICKLE3* | blood | 66-70 | M | n/a | n/a | no |
| 257 | Control group for *PRICKLE3* | blood | 71-75 | M | n/a | n/a | no |

| | | | | | | | |
|---|---|---|---|---|---|---|---|
| 258 | Control group for *PRICKLE3* | blood | n/a | F | n/a | n/a | no |
| 259 | Control group for *PRICKLE3* | blood | n/a | M | n/a | n/a | no |
| 260 | Control group for *PRICKLE3* | blood | 61-65 | M | n/a | n/a | no |
| 261 | Control group for *PRICKLE3* | blood | n/a | F | n/a | n/a | no |
| 262 | Control group for *PRICKLE3* | blood | 71-75 | F | n/a | n/a | no |

[a] Exact ages have been replaced with non-overlapping age ranges to comply with data de-identification requirements. All age-related statistical analyses were performed using exact age values.

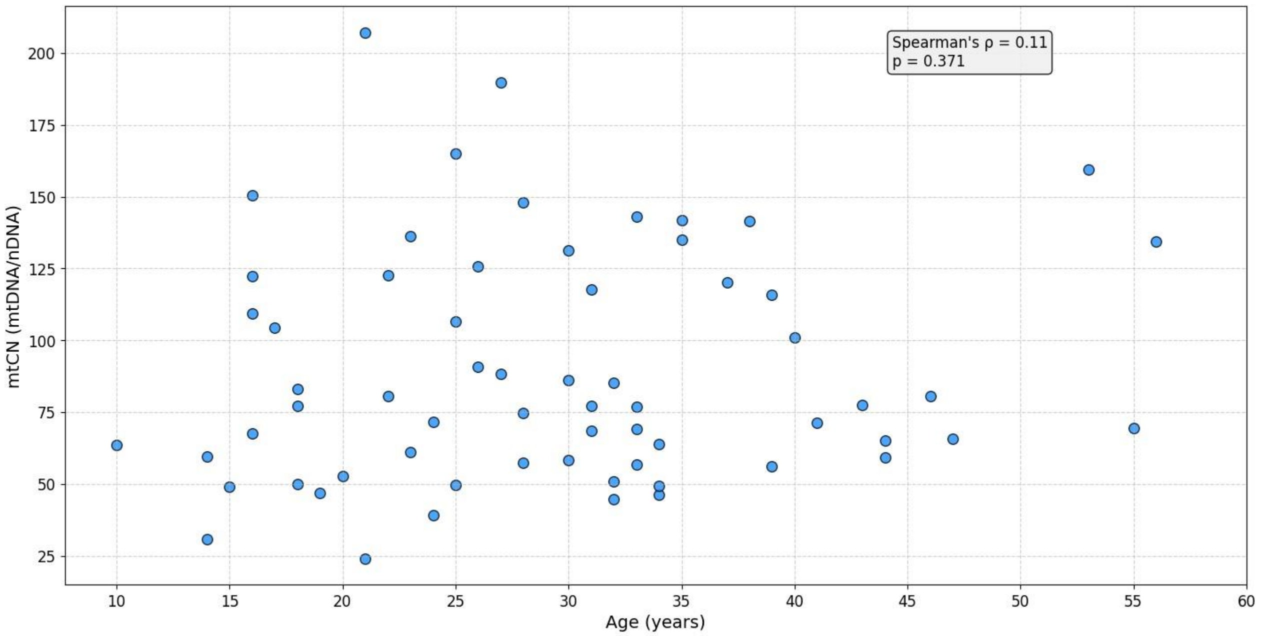


**Fig. S1** A scatter plot illustrating the relationship between the approximate age of LHON symptom onset and mtCN in symptomatic carriers. Spearman's rank correlation test revealed a correlation coefficient ρ of 0.114 with a *p*-value of 0.371, indicating no statistically significant association
